# Shifting of economic responsibilities on vulnerable shoulders for survival of families: a scenario from mining industry in India

**DOI:** 10.1101/19012591

**Authors:** Pankaja Raghav, Abhishek Jaiswal, Manoj Kumar Gupta, Saranaya Rajavel, Diksha Dhupar

## Abstract

**Context:** Mining industry has boomed precipitously to provide for demand of metal and minerals for emerging infrastructure. It is major occupation in India involving large work force, although it is hazardous. Aims: To assess the socio-demographic profile of sandstone mine workers in Jodhpur and to identify their major social issues.

**Material and Methods:** Study design: Cross-sectional study; Setting: Residential area of mine workers, Jodhpur, Rajasthan; Participants: Mine workers residing in residential areas in Jodhpur district for more than 6 months, and who gave consent for participation in study. Data on socioeconomic status and occupation of each family members was collected; Analysis: Data was analysed using SPSS v.23. P value <0.05 was considered as statistically significant.

**Results:** One thousand six hundred four mine workers were enrolled in study. More than three fourth (78.7%) of them were male. Their age ranged between 14 to 82 years with mean age of 37.62 ± 12.30 years. Mean age of female workers was significantly higher than of male workers. Average years of working for males was significantly higher than females. Mean age of entry into mines for male workers (20.33 ± 6.69) was significantly lower than females (24.59 ± 8.52). About 2.68% children below 14 years of age were involved in any sort of income generation activity. Conclusions: Majority of mine workers were of young age group. Major socioeconomic issues found were illiteracy, child marriage, child labour, loneliness and shifting economic responsibilities on females.

**Strengths and limitations:** Robust dataset, complete enumeration, large sample size.

**Key messages:** *What is already known about this subject?:* The miners work under exploitative conditions and their families live in severe poverty with no access to health services. Families living close to the mines and are prone to developing silicosis from secondary exposure, they are also prone for malnourishment, tuberculosis and other comorbid conditions. Early deaths of the male earning member lead to vulnerable members of family to take the job for their livelihood. There are many reported widow villages where there is no male member due to death of the same by pneumoconiosis.

*What is the new findings?:* There is increasing burden on the shoulders of the vulnerable groups (children, females and elderly) among families of sandstone miners in India. Illiteracy, child marriage, child labour and loneliness are among major social problems in these families.

*How might this impact on policy in the foreseeable future?:* Families where the main bread earner died of pneumoconiosis could be provided pension to supplement for their food and health expenses requirement. Rajasthan state in India has recently launched “Silicosis Policy”, and this study signifies the importance of implementing such polices in other states in India.

## Introduction

Importance of mining has increased precipitously to provide for the demand of metal and minerals for the emerging infrastructure.^1^ Although mining is a hazardous occupation, yet it is one of the major occupations in India involving large number of work force which is going to grow in future. ^2,3^

Approximately 90% sandstone in India is acquired from the land of the state of Rajasthan which is largest state of India. This is used in India as well as exported to Canada, Japan, and countries in the Middle East.^4^ There are near about 30,000 mines in Rajasthan, in which more than 2.5 million people are employed. Out of those mines around 1300 are sandstone mines, in which more than 71,000 miners are working.^4^ Those workers are involved in various activities in mines like; drilling, blasting, crushing the large stone to smaller one and loading-unloading of slab from trucks, that expose them to high levels of silica dust for prolonged periods of time.^1^ The geological configuration of Jodhpur district of Rajasthan is interesting and is represented by rocks like; sandstone, limestone, granite, rhyolite, schist, phyllite, and slate.^5^ It shares a common border with five districts: Bikaner and Jaisalmer in the north and north-west, respectively; Banner and Pali in the south-west and south-east, respectively; and Nagaur in the northeast. Jodhpur is the zonal headquarter to controls all mineral-related activities in western part of Rajasthan.

Although there are studies carried out on mine workers in Jodhpur but they are focused on disease aspects (silicosis).^6-8^ It has been proved that mine workers, especially males are usually exposed to silica since childhood, and die due to silicosis at quite early age.^9^ After their death somebody from their dependent family members, who in most of the cases are either their widows or young children, is bound to work in mines for their survival. And this cycle is continuing over generations. There is deficiency of robust datasets to prove these facts, henceforth this study was carried out with the aim to assess the socio-demographic profile of sandstone mine workers in Jodhpur and to identify their major social issues.

## Material and Methods

This was a descriptive cross-sectional study which was conducted for one year (April 2018 to March 2019) in Jodhpur district of Rajasthan state. Study was carried out in residential area of mine workers. Mine owners of all the selected mines were contacted and explained about the purpose of the study. All the mine workers who were present at the time of survey were included in the study. After establishing the rapport with mine owners they were informed about the study, its objective and the questions we were going to ask. Informed consent was taken. Participants who gave consent for the study were asked questions on socioeconomic status and occupation of each family members. The study was approved by Institutes Ethical Committee of All India Institute of Medical Sciences (AIIMS), Jodhpur. Data was analysed using SPSS v.23, appropriate tables and graphs were prepared and inferences were drawn using chi-square and t-test. P value <0.05 was considered as statistically significant.

### Patient and Public involvement

Just after the conception of idea of this study the investigator talked to the resident mine workers regarding the study and how it will be conducted, the information that will be asked and the benefit of the study. Mine workers were not involved in the design of study/ framing of the research question/ conduct of the study. The study findings were discussed with the mine-workers verbally and appropriate advise was given.

## Results

Table 1 depicts that a total of 1604 mine workers were enrolled in the study. More than three fourth (78.7%) of them were male. Their age ranged between 14 to 82 years with mean age of 37.62 ± 12.30 years. The data was normally distributed according to age of the mine workers (Figure 1). Most of the workers (78%) were in the age group of 20-49 years. Thirty-nine workers were below 20 years of age and 7.1% workers were elderly. Mean age of female workers was significantly higher than the mean age of male workers. Majority (93.7%) of them were married. The proportion of widows was significantly higher among female workers, while proportion of unmarried was significantly higher among males. It was affirmed that 13.22% mine workers got married below the legal age of marriage (21 for male and 18 for female). This proportion was significantly higher for females. More than half of the mine workers were illiterate (53.4%), and this status of illiteracy was significantly higher among females.

**Table 1:** Socio demographic attributes of mine workers (N=1604)

|  | Male (n=1263) |  | Female (n=341) |  | Total |  | p value |
| --- | --- | --- | --- | --- | --- | --- | --- |
|  | n | % | N | % | n | % |  |
| **Age (years)** |  |  |  |  |  |  |  |
| <20 | 34 | 2.70 | 5 | 1.50 | 39 | 2.40 | 0.12 |
| 20-29 | 331 | 26.20 | 78 | 22.90 | 409 | 25.50 |  |
| 30-39 | 367 | 29.10 | 105 | 30.80 | 472 | 29.40 |  |
| 40-49 | 295 | 23.40 | 74 | 21.70 | 369 | 23.00 |  |
| 50-59 | 145 | 11.50 | 56 | 16.40 | 201 | 12.50 |  |
| ≥60 | 91 | 7.20 | 23 | 6.70 | 114 | 7.10 |  |
| **Mean Age** | 37.62±12.30 |  | 39.16±12.56 |  | 37.95±12.37 |  | **0.041** |
| **Age range** | 68 (14-82) |  | 57 (18-75) |  | 68 (14-82) |  |  |
| **Marital Status** |  |  |  |  |  |  |  |
| Unmarried | 66 | 5.20 | 3 | 0.90 | 69 | 4.30 | <0.05 |
| Married | 1186 | 93.90 | 317 | 93.00 | 1503 | 93.70 |  |
| Widow | 11 | 0.90 | 21 | 6.20 | 32 | 2.0 |  |
| **Married before legal age** | 127 | 10.05 | 85 | 24.92 | 212 | 13.22 | **<0.05** |
| **Educational Status** |  |  |  |  |  |  |  |
| Illiterate | 562 | 44.50 | 294 | 86.20 | 856 | 53.40 |  |
| Pre Primary | 161 | 12.70 | 15 | 4.40 | 176 | 11.00 |  |
| Primary | 220 | 17.40 | 22 | 6.50 | 242 | 15.10 |  |
| Middle | 200 | 15.80 | 8 | 2.30 | 208 | 13.00 | **<0.05** |
| Secondary | 75 | 5.90 | 1 | 0.30 | 76 | 4.70 |  |
| Higher Secondary | 35 | 2.80 | 0 | 0.00 | 35 | 2.20 |  |
| Graduate and above | 10 | 0.80 | 1 | 0.30 | 11 | 0.70 |  |

It is evident from table 2 that about two third (62.6%) of the workers were working for more than 10 years in mines. The mean period of work was 16.71 ± 9.72 years and ranged between 1 year to 60 years. Average years of working for males was significantly higher than females. The mean age of entry into mines for male workers (20.33 ± 6.69) was significantly lower than female workers (24.59 ± 8.52). Workers started working in mines from 9 years of age to 62 years of age.

**Table 2:**
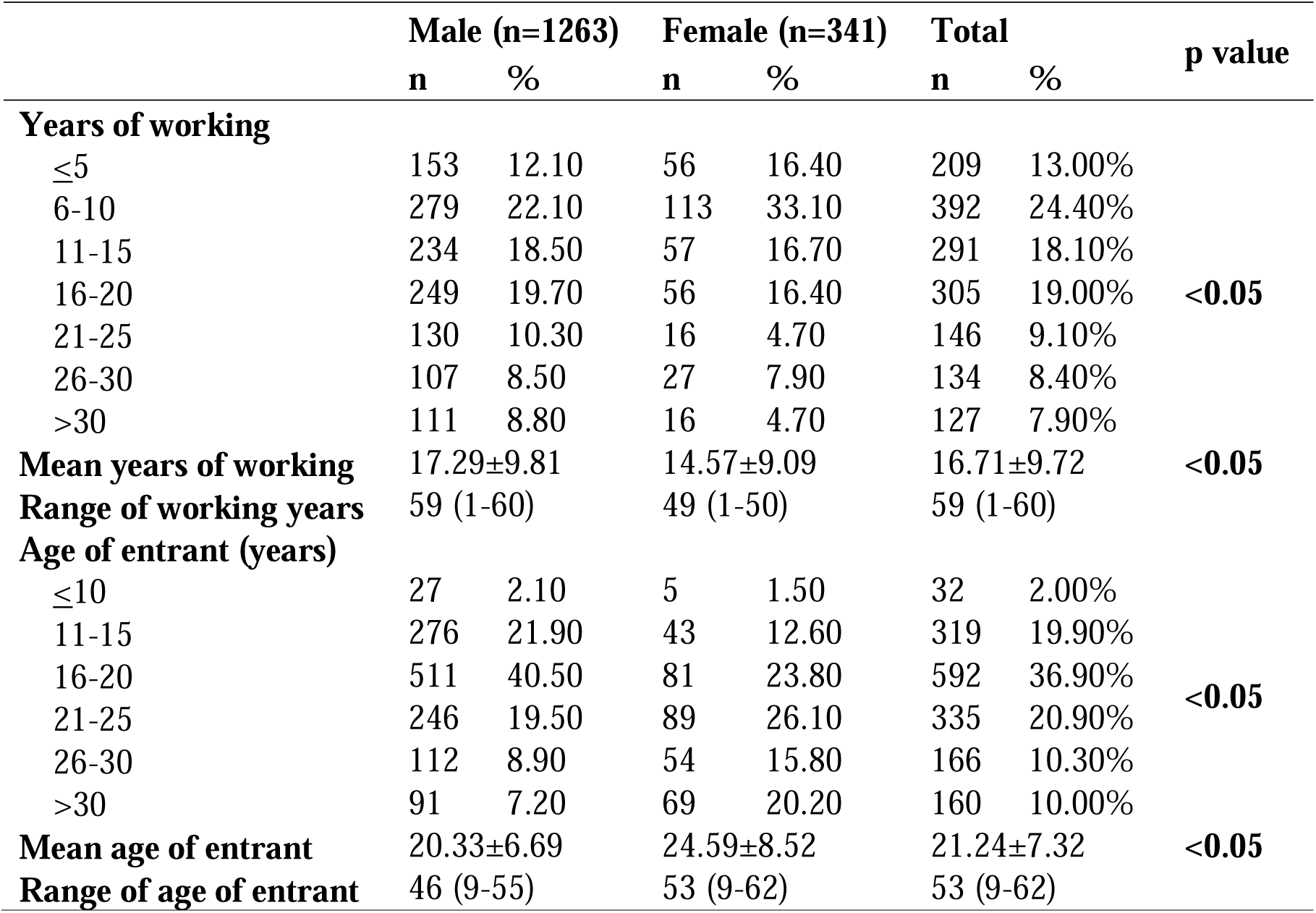
Work related attributes of mine workers (N=1604)

|  | Male (n=1263) |  | Female (n=341) |  | Total |  | p value |
| --- | --- | --- | --- | --- | --- | --- | --- |
|  | n | % | n | % | n | % |  |
| **Years of working** |  |  |  |  |  |  |  |
| ≤5 | 153 | 12.10 | 56 | 16.40 | 209 | 13.00% |  |
| 6-10 | 279 | 22.10 | 113 | 33.10 | 392 | 24.40% |  |
| 11-15 | 234 | 18.50 | 57 | 16.70 | 291 | 18.10% |  |
| 16-20 | 249 | 19.70 | 56 | 16.40 | 305 | 19.00% | **<0.05** |
| 21-25 | 130 | 10.30 | 16 | 4.70 | 146 | 9.10% |  |
| 26-30 | 107 | 8.50 | 27 | 7.90 | 134 | 8.40% |  |
| >30 | 111 | 8.80 | 16 | 4.70 | 127 | 7.90% |  |
| **Mean years of working** | 17.29±9.81 |  | 14.57±9.09 |  | 16.71±9.72 |  | **<0.05** |
| **Range of working years** | 59 (1-60) |  | 49 (1-50) |  | 59 (1-60) |  |  |
| **Age of entrant (years)** |  |  |  |  |  |  |  |
| ≤10 | 27 | 2.10 | 5 | 1.50 | 32 | 2.00% |  |
| 11-15 | 276 | 21.90 | 43 | 12.60 | 319 | 19.90% |  |
| 16-20 | 511 | 40.50 | 81 | 23.80 | 592 | 36.90% | **<0.05** |
| 21-25 | 246 | 19.50 | 89 | 26.10 | 335 | 20.90% |  |
| 26-30 | 112 | 8.90 | 54 | 15.80 | 166 | 10.30% |  |
| >30 | 91 | 7.20 | 69 | 20.20 | 160 | 10.00% |  |
| **Mean age of entrant** | 20.33±6.69 |  | 24.59±8.52 |  | 21.24±7.32 |  | **<0.05** |
| **Range of age of entrant** | 46 (9-55) |  | 53 (9-62) |  | 53 (9-62) |  |  |

Table 3 depicts that 110 (6.9%) mine workers were living alone. As much as 124 (7.7%) workers were having only one family member in their house. It is evident from figure 2 and 3 that, the other family member for majority (82.1%) of the male workers was their wife followed by parents (8.4%), brother (4.2%), son/grandson (3.1%) and daughter (2.1%). While for most of the female miners (45%) the other family member was son/grandson followed by husband (38%), daughter (14%) and mother in law (3%). Average family size of the mine workers was 4.45 ± 1.57 which was ranging from 1 to 11. This average family size was significantly large for male workers (4.87±1.93) as compared to female workers (4.23±1.98). Almost 40% of the mine workers were having at least one more family member as mine worker.

**Table 3:** Family attributes of mine workers according to gender distribution (N=1604)

|  | Male (n=1263) |  | Female (n=341) |  | Total |  | p value |
| --- | --- | --- | --- | --- | --- | --- | --- |
|  | n | % | n | % | n | % |  |
| **Family size** |  |  |  |  |  |  |  |
| Single | 70 | 5.50 | 40 | 11.70 | 110 | 6.90 | **<0.05** |
| Two | 95 | 7.50 | 29 | 8.50 | 124 | 7.70 |  |
| Three | 132 | 10.50 | 53 | 15.50 | 185 | 11.50 |  |
| Four | 210 | 16.60 | 63 | 18.50 | 273 | 17.00 |  |
| Five | 260 | 20.60 | 74 | 21.70 | 334 | 20.80 |  |
| More than five | 496 | 39.30 | 82 | 24.00 | 578 | 36.00 |  |
| Mean family size | 4.87±1.93 |  | 4.23±1.98 |  | 4.45±1.57 |  | <0.05 |
| Range of family size | 10 (1-11) |  | 8 (1-9) |  | 10 (1-11) |  |  |
| Number of mine workers among family members* |  |  |  |  |  |  |  |
| None | 809 | 67.80% | 96 | 31.90% | 905 | 60.60% |  |
| One | 293 | 24.60% | 136 | 45.20% | 429 | 28.70% | <0.05 |
| Two or more | 91 | 7.60% | 69 | 22.90% | 160 | 10.70% |  |

\*Proportion has been calculated out of those who were living with at least one family member

Table 4 illustrates the age and sex distribution of family members of mine workers.

**Table 4:** Age and sex distribution of family members of mine workers.

| **No. of family members in different age groups** | **Male** |  | **Female** |  | **Total** |  | **p value** |
| --- | --- | --- | --- | --- | --- | --- | --- |
|  | **n** | **%** | **n** | **%** | **n** | **%** |  |
| Up to 5 years | 440 | 10.70 | 403 | 11.60 | 843 | 11.10 |  |
| Children 6-9 years | 418 | 10.20 | 375 | 10.80 | 793 | 10.40 |  |
| Adolescents (10-19 years) | 1076 | 26.20 | 913 | 26.20 | 1989 | 26.20 | <0.05 |
| Adults (20-59 years) | 1974 | 48.10 | 1698 | 48.70 | 3672 | 48.40 |  |
| Geriatric (≥60 years) | 199 | 4.80 | 98 | 2.80 | 297 | 3.90 |  |
| **Total** | **4107** | **100.00** | **3487** | **100.00** | **7594** | **100.00** |  |

Table 5 depicts that 2.68% children below 14 year of age were involved in any sort of income generation activity. This child labour was significantly more prevalent among male children. The youngest child who was labour was of 6 years old. Youngest mine workers were two, and both were 7 years old. Besides that, 6.5% children between age 14 to 18 year of age were mine workers, which is a hazardous occupation.

**Table 5:** Status of child labour in families of mine workers.

| Occupation | Male<br>n | % | Female<br>n | % | Total<br>n | % | p value |
| --- | --- | --- | --- | --- | --- | --- | --- |
| <14 years |  |  |  |  |  |  |  |
| Farmer | 2 | 0.20 | 3 | 0.30 | 5 | 0.20 | 0.034 |
| Labour | 27 | 2.10 | 21 | 1.80 | 48 | 1.90 |  |
| Mine Worker | 12 | 0.90 | 2 | 0.20 | 14 | 0.60 |  |
| Unemployed | 772 | 58.80 | 746 | 63.00 | 1518 | 60.70 |  |
| Student | 501 | 38.10 | 413 | 34.90 | 914 | 36.60 |  |
| Total | 1314 | 100.00 | 1185 | 100.00 | 2499 | 100.00 |  |
| 14 to 18 years |  |  |  |  |  |  |  |
| Farmer | 7 | 1.30 | 4 | 0.90 | 11 | 1.10 | <0.05 |
| Labour | 50 | 9.30 | 15 | 3.50 | 65 | 6.70 |  |
| Mine Worker | 49 | 9.10 | 14 | 3.30 | 63 | 6.50 |  |
| Unem | 163 | 30.40 | 197 | 46.20 | 360 | 37.40 |  |
| Student | 268 | 49.90 | 196 | 46.00 | 464 | 48.20 |  |
| Total | 537 | 100.00 | 426 | 100.00 | 963 | 100.00 |  |

## Discussion

After extensive literature search, it was observed that there is lack of epidemiological studies which describes the socio-demographic profiling of sandstone mine workers in India and highlight their social issues. This study was conducted to fulfil this knowledge gap.

### Demographic and occupational profiling of mine workers

In this study, about three fourth of the mine workers were males. Similar pattern of gender distribution of mine workers has been reported by Shamim M. et.al. (2017) in Rajasthan and Tiwari RR (2010) in Gujarat.^10,11^ Different gender distribution (55.1% males and 44.9% females) has been observed by Tiwari RR (2005) among quartz stone workers.^12^

In the present study, about two third (62.6%) of the workers were working in mines for more than 10 years. Nandi S et.al. (2018) has also reported similar level of work experience among mine workers.^3^ Contrary to this Rajashekar S et al. (2017) has reported that majority of the mine workers in Karnataka had total work experience of five years.^13^ The mean years of working in the present study was 16.71 ± 9.72 years which is lower than the mean years of work reported by Ahmad A (2015)^14^ and Shamim M et al. (2017).^11^ Mine workers were involved in various activities in the mines from drilling, blasting, cutting of stones to carrying the stones, loading the stones in the vehicles, cleaning of the waste material.

### Social issues of mine workers

In this study, following major social issues were identified among mine workers and their family members.

#### 1. Illiteracy

More than half of the mine workers were illiterate in the present study. Higher level of illiteracy among mine workers in North India has also been reported by other studies.^10,14,15^ Mine workers of Southern part of India had relatively better educational status.^13^ The proportion of illiteracy was significantly higher among the female workers compared to the male workers. Study also highlights that among family members of mine workers, more than half of the children between 14 to 18 year age group were dropped out from the schools. This proportion of dropouts was significantly more for girls, which is opposite to the findings reported by Government of India.^16^

#### 2. Child marriage

India has maintained laws against child marriage since 1929, but still child marriage is quite prevalent in certain areas of the country.^17,18^ Prevalent and deep routed social norms pushes children and especially girls for early marriage and discourages education.^19^ It was observed that 13.22% mine workers got married below the legal age of marriage (21 for male and 18 for female) in the present study, and this proportion was significantly higher among the female workers. Girls are bounded by social norms which force them for early marriage and discourage education.^19^ This figure is quite lower than the proportion reported by National Family Heath Survey 4^th^ round (NFHS IV) for both males and females for rural Rajasthan.^20^

#### 3. Loneliness

Loneliness is considered as a disease. It may lead to a decline of well-being, depression, suicidal behaviour, sleep problems, disturbed appetite etc.^21^ A strong need of the specific public support systems and health care strategies to take care of these people has already been emphasised.^22,23^ Although average family size of the mine workers in this study was 4.45 ± 1.57, which is in accordance with the average family size reported in NFHS IV ^24^, about 7% of the mine workers were living alone in their houses. Relatively larger average household size for mine workers was reported by Ahmad A (2014 &2017).^14,25^

#### 4. Female mine workers

In the present study, the mean age of the female mine workers (39.16 ± 12.56 years) was significantly higher than the male workers (37.62 ± 12.30 years). This could be due to late age of entry into the mining work, as evident by the significantly higher age of entrant in female workers (24.59 ± 8.52 years) in mines as compared to the male workers (20.33 ± 6.69 years). This higher mean age of entry into mine for females may be due to the reason that females start working in mines to fulfil their financial needs only after marriage, when their husband got diseased or died due to silicosis and they have several children to care for.^26^ This statement can be supported by the fact that the proportion of widows was significantly higher among female workers (6.4%), compared to male workers (0.2%), and mean years of working was significantly lower among female workers (14.57 ± 9.09) compared to male workers (17.29 ± 9.81). Among two-member family of male mine workers, the other member who was living with them was wife for majority (82.1%), while husbands as other family member was with only 38% of the female mine workers living in two-member family. This also supports the finding that wives join the work after the husband could no longer support them because of illness/death.

Significant strides have been taken by Government of Rajasthan in this direction by launching the Silicosis Policy in October 2019. As per this policy, widow of a mine workers died due to silicosis, will get all the social security benefits from the state government including a monthly pension of up to Rs. 1500.^27^

On the other side of the coin, the mean age of entry into the mining occupation for males may be lower because generation wise these families are following the patriarchal culture of working in mines. So, the male members start working at early age to earn for their family, giving very less priority to education. Sometimes the carry forwarded loans taken by their ancestors also forced male members to start work in mines at early age.^25^

#### 5. Child Labour

Child labour is physically, mentally, socially and morally dangerous and harmful to children, and also negatively relates to school attendance and learning.^28,29^ Although there is provision of free and compulsory education for children between 6 to 14 years of age, and prohibition of employment of children between 14 to 18 years of age in hazardous occupations in Constitution of India, yet child labour is quite prevalent in India.^30^ Children are forced into labor and sometimes in hazardous occupations due to poverty and to pay family debt.^31,32^ Child labour was quite prevalent in the study area. This finding is well supported by the prevalence of child labour reported in 2011 census survey of the country.^30^ A substantial contribution of child labour force among mine workers has also been highlighted by Murlidhar V. (2015).^9^ The existence of child labor in the mining industry is to supplement the earning of parents.^33^ The other reason can be the premature death of bread earner due to silicosis.^34^

#### 6. Elderly mine workers

In the present study, most of the mine workers (78%) were in the age group of 20-49 years. This corresponds to the age groups of miners observed by Solanki J et.al. (2014) and Nandi S et.al. (2018) in Jodhpur district.^35,36^ Mine workers were belonging to almost all age groups (adolescents to elderly) with mean age of 37.62 ± 12.30 years. Mining is a hazardous occupation and is not considered safe workplace for elderly people. As much as 7.1% mine workers were elderly in the present study. These findings are in accordance to the findings reported by other studies.^4,37^

## Conclusion

In the present study, it was found that most of the mine workers were in young age group and predominantly males. Major social issues among mine workers were illiteracy, child marriage, child labour, loneliness and shifting economic responsibilities on females. Based on the findings it can be concluded that, economic responsibilities are shifting on vulnerable shoulders (children, females and elderly) in sandstone mines in India.

## Data Availability

Data are available upon reasonable request

## Acknowledgement

Authors acknowledge the mine workers for taking part in the study.

## References

1. Detection of Silicosis among stone mine workers from Karauli district [Internet]. Nagpur: National Institute of Miners Health (NIMH); 2011. Available from: http://aravali.org.in/themes/upload/files/276725.pdf

2. Cho KS, Lee SH. Occupational health hazards of mine workers. Bull World Health Organ. 1978;56(2):205–18.

3. Nandi SS, Dhatrak SV, Chaterjee DM, Dhumne UL. Health Survey in Gypsum Mines in India. Indian J Community Med Off Publ Indian Assoc Prev Soc Med. 2009 Oct;34(4):343–5.

4. Ahmad A. Silicosis, Mining, and Occupational Health in India’s Sandstone Industry - EHS Journal [Internet]. 2015 [cited 2019 Jul 11]. Available from: http:/ehsjournal.org/absar-ahmad/silicosis-mining-and-occupational-health-in-indias-sandstone-industry/2015/

5. Solanki J, Gupta S, Chand S. Oral Health of Stone Mine Workers of Jodhpur City, Rajasthan, India. Saf Health Work. 2014 Sep;5(3):136–9.

6. Shafiei M, Ghasemian A, Eslami M, Nojoomi F, Rajabi-Vardanjani H. Risk factors and control strategies for silicotuberculosis as an occupational disease. New Microbes New Infect. 2018 Nov 15;27:75–7.

7. Nandi S, Burnase N, Barapatre A, Gulhane P, Dhatrak S. Assessment of Silicosis Awareness among Stone Mine Workers of Rajasthan State. Indian J Occup Environ Med. 2018;22(2):97–100.

8. Singh SK, Chowdhary GR, Purohit G. Assessment of impact of high particulate concentration on peak expiratory flow rate of lungs of sand stone quarry workers. Int J Environ Res Public Health. 2006 Dec;3(4):355–9.

9. Murlidhar V. An 11-year-old boy with silico-tuberculosis attributable to secondary exposure to sandstone mining in central India. BMJ Case Rep [Internet]. 2015 Jun 23 [cited 2019 Oct 7];2015. Available from: https://www.ncbi.nlm.nih.gov/pmc/articles/PMC4480122/

10. Tiwari RR, Narain R, Sharma YK, Kumar S. Comparison of respiratory morbidity between present and ex-workers of quartz crushing units: Healthy workers’ effect. Indian J Occup Environ Med. 2010;14(3):87–90.

11. Shamim M, Dr Waheeb D.M. Alharbi, Dr Tariq Sultan Pasha, Dr Mohamed Osama Mustafa Nour. Silicosis, A Monumental Occupational Health Crisis In Rajasthan-An Epidemiological Survey. 2017 Aug 10 [cited 2019 Jul 17]; Available from: https://zenodo.org/record/841120

12. Tiwari RR, Sharma YK, Saiyed HN. Peak expiratory flow and respiratory morbidity: a study among silica-exposed workers in India. Arch Med Res. 2005 Apr;36(2):171–4.

13. Rajashekar S, Sharma P. Morbidity among mine workers: a cross sectional study in Chitradurga, Karnataka, India. Int J Community Med Public Health. 2017 Jan 25;4(2):378–84.

14. Ahmad A. A study of Miners, Demographics and Health Status in Jodhpur District of Rajasthan, India. Int J Dev Stud Res. 2014;3(1):113–21.

15. Yadav SP, Anand PK, Singh H. Awareness and Practices about Silicosis among the Sandstone Quarry Workers in Desert Ecology of Jodhpur, Rajasthan, India. J Hum Ecol. 2011 Mar 1;33(3):191–6.

16. Annual average drop out rate of girls is lesser than drop out rate of boys at secondary level | Government of India, Ministry of Human Resource Development [Internet]. [cited 2019 Oct 12]. Available from: https://mhrd.gov.in/annual-average-drop-out-rate-girls-lesser-drop-out-rate-boys-secondary-level

17. Prevalence of Child Marriage and its Impact on the Fertility and Fertility Control Behaviors of Young Women in India [Internet]. [cited 2019 Oct 12]. Available from: https://www.ncbi.nlm.nih.gov/pmc/articles/PMC2759702/

18. Chandra-Mouli V, Plesons M, Barua A, Sreenath P, Mehra S. How can collective action between government sectors to prevent child marriage be operationalized? Evidence from a post-hoc evaluation of an intervention in Jamui, Bihar and Sawai Madhopur, Rajasthan in India. Reprod Health. 2018 Jun 28;15(1):118.

19. Students and brides: a qualitative analysis of the relationship between girls’ education and early marriage in Ethiopia and India [Internet]. [cited 2019 Oct 12]. Available from: https://www.ncbi.nlm.nih.gov/pmc/articles/PMC6322316/#CR18

20. MOHFW. National Family Health Survey IV Rajasthan Fact Sheet [Internet]. 2015. Available from: http://rchiips.org/nfhs/pdf/NFHS4/RJ_FactSheet.pdf

21. Tiwari SC. Loneliness: A disease? Indian J Psychiatry. 2013 Dec;55(4):320.

22. Agrawal S. Effect of Living Arrangement on the Health Status of Elderly in India: Findings from a national cross sectional survey. Asian Popul Stud. 2012 Mar;8(1):87–101.

23. Goswami S, Deshmukh PR. How “Elderly Staying Alone” Cope Up with their Age and Deteriorating Health: A Qualitative Exploration from Rural Wardha, Central India. Indian J Palliat Care. 2018;24(4):465–71.

24. MOHFW. National Family Health Survey IV [Internet]. 2015. Available from: http://rchiips.org/NFHS/NFHS-4Reports/India.pdf

25. Ahmad A. Socio-economic and health status of sandstone miners: a case study of Sorya village, Karauli, Rajasthan. Int J Res Med Sci. 2017 Jan 8;3(5):1159–64.

26. Tears of Dust, A study of women labourers in the mines of Makrana and Jodhpur, GRAVIS, Jodhpur, December 1995.

27. Rajasthan government launches silicosis policy for mining workers | Jaipur News - Times of India [Internet]. The Times of India. [cited 2019 Oct 11]. Available from: https://timesofindia.indiatimes.com/city/jaipur/raj-govt-launches-silicosis-policy-for-mining-workers/articleshow/71431702.cms

28. Ibrahim A, Abdalla SM, Jafer M, Abdelgadir J, de Vries N. Child labor and health: a systematic literature review of the impacts of child labor on child’s health in low- and middle-income countries. J Public Health Oxf Engl. 2019 Mar;41(1):18–26.

29. Putnick DL, Bornstein MH. Is Child Labor a Barrier to School Enrollment in Low- and Middle-Income Countries? Int J Educ Dev. 2015 Mar 1;41:112–20.

30. FACT Sheet: Child labour in India [Internet]. 2017 [cited 2019 Oct 7]. Available from: http://www.ilo.org/newdelhi/whatwedo/publications/WCMS_557089/lang--en/index.htm

31. Srivastava K. Child labour issues and challenges. Ind Psychiatry J. 2011;20(1):1–3.

32. Radfar A, Asgharzadeh SAA, Quesada F, Filip I. Challenges and perspectives of child labor. Ind Psychiatry J. 2018;27(1):17–20.

33. Sinha SC. The Unorganised Sector in India: Extending the Debate to Mining and Quarrying - Seminar organised by Institute of Development Studies (IDS), Jaipur, [Internet]. 2013 [cited 2019 Jul 17]. Available from: https://www.indiawaterportal.org/events/call-papers-unorganised-sector-india-extending-debate-mining-and-quarrying-seminar-organised

34. Ahmad A. A study of sandstone miner: notes from the field. Int J Med Sci Public Health. 2015;4(3):433.

35. Solanki J, Gupta S, Chand S. Oral Health of Stone Mine Workers of Jodhpur City, Rajasthan, India. Saf Health Work. 2014 Sep;5(3):136–9.

36. Nandi S, Burnase N, Barapatre A, Gulhane P, Dhatrak S. Assessment of Silicosis Awareness among Stone Mine Workers of Rajasthan State. Indian J Occup Environ Med. 2018;22(2):97–100.

37. Verma S, Chaudhari S. Safety of Workers in Indian Mines: Study, Analysis, and Prediction. Saf Health Work. 2017 Sep;8(3):267–75.

